# Assessment of the Usability of SARS-CoV-2 Self Tests in a Peer-Assisted Model among Factory Workers in Bengaluru, India

**DOI:** 10.1101/2023.11.20.23298784

**Authors:** Meghana Ratna Pydi, Petra Stankard, Neha Parikh, Purnima Ranawat, Ravneet Kaur, AG Shankar, Angela Chaudhuri, Sonjelle Shilton, Aditi Srinivasan, Joyita Chowdhury, Elena Ivanova Reipold

**Affiliations:** Swasti - The Health Catalyst, Bengaluru, Karnataka, India; FIND, Geneva, Switzerland

**Keywords:** Usability of self-testing, Sars-CoV-2 testing, Acceptability of self-testing, peer-assisted self-testing, Workplace COVID-19 testing, Pandemic preparedness, Community-based self-testing, Self-testing for marginalized populations

## Abstract

In order to mitigate the inequities in health outcomes and healthcare access for vulnerable populations during the COVID-19 pandemic, the government of India introduced antigen-based SARS-CoV-2 self-testing kits for self-administered use. In this study, we aimed to determine the usability of these nasal-sampling-based self-tests in a peer-assisted model among factory workers in Bengaluru. The mixed-method cross-sectional study was conducted with 106 factory workers, spanning two sites from February to March 2022 in Bengaluru, India. Panbio™ COVID-19 Antigen Self-Test kit and the mobile application NAVICA for self-reporting results were used. A peer assistant distributed test kits, guided participants on conducting tests and using the app, and offered demonstrations with their own kit, ensuring no contact with the participants’ kits. Findings were encapsulated by an observer, who used standardized product-specific usability checklists and pictures of contrived results to assess the usability of the kit and mobile application, result interpretation, and the efficiency of peer instruction/demonstration. Additionally, a post-test survey and focus group discussions with selected participants and peer assistants were conducted to understand user perceptions of the facilitators and barriers to usability. Study findings show that the overall usability score of the test kit with peer assistance was 75.9%, rising to 80.7% for critical steps and 33.8% for all critical steps in uploading results through NAVICA. Additionally, it was seen that peer assistants provided accurate instructions and support for 93.4% of the tests. Among the critical steps in test kit use, maximum errors were made in sample collection and using the correct amount of buffer solution. Concordance between the participant and observer/NAVICA was 97.9%. 62.0% and 56.6% of the participants reported confidence in a) performing and interpreting the test and b) capturing and uploading their results using the mobile application with the assistance of a peer, respectively. Less than half the participants reported confidence in performing these steps independently. The study indicates that the COVID-19 nasal self-testing kit has good usability in factories’ peer-assisted workplace testing model. Such models can empower vulnerable worker groups to access early detection and self-care tools equitably.

## Introduction

Coronavirus disease 2019 (COVID-19), caused by the severe acute respiratory syndrome coronavirus-2 (SARS-CoV-2), took severe health, social and economic toll worldwide. As of July 2023, India reported 44 million cases and 531,913 deaths ^(1)^.

India’s manufacturing sector contributes 16% to the country’s gross domestic product (GDP) and employs about 20% of the country’s workforce. During the COVID-19 pandemic, the sector was heavily impacted. Severe workforce shortages and factory shutdowns negatively affected employee turnover and revenue. Between 2020-1, the Index of Industrial Production decreased by 9.6%, reflecting setbacks to core manufacturing during the pandemic’s second wave ^(2)^.

The workforce in manufacturing industries consists of semi-formal or informal factory workers, including migrants and daily wage earners ^(3)^, typically living in informal settlements that define their access to water and sanitation, and healthcare ^(4)^. The industrial slowdown directly impacted the vulnerable workers. Karnataka, which accounted for 5.7% of the total registered factories in 2017-8 in India, saw a permanent shutdown of 754 factories, with nearly 46000 workers losing their jobs since the onset of the pandemic between 2020 and 2021 ^(5)^ Recent studies found high unemployment and reduced incomes in Bengaluru, as a result of the pandemic.

Factory workers are also at increased risk of infections and poor health outcomes, including COVID-19-related morbidity and mortality, due to working in close proximity for long periods. Vaccination policies restricting access to those below 45 years of age and vaccine hesitancy among factory workers further exacerbated their vulnerability. In addition, most factory workers are migrants and face issues accessing local services due to language barriers. Low awareness of COVID-19 testing, vaccination services, poor understanding of transmission prevention, limited perception of risk, and fear of discrimination following disclosure of a COVID-19 positive status further increase risk of infection, limit testing and accurate diagnosis^(6)^.

As the second wave of infections eased after June 2021, businesses re-opened and implemented occupational safety programs to prevent outbreaks and support employees that tested positive. These were mandated for large workplaces by state governments. ^(7,8)^. ￼Successful implementation of these programs required the availability of timely diagnostic testing to facilitate case identification and quarantine. However, the supply of Reverse Transcription - Polymerase Chain Reaction (RT-PCR) testing for SARS-CoV-2 in health facilities was constrained by the need for significant laboratory capacity and trained clinical staff, as well as a shortage of molecular reagents, supplies, and equipment. Fear of painful testing procedures and long waiting and travel times due to limited testing facilities further constrained demand ^(9,10,11)^. ￼￼

Rapid Antigen Testing (RAT) for COVID-19 has been recommended and successfully implemented globally in workplaces and clinical settings ^(12,13)^. It improves individuals’ access to testing and, due to shorter turnaround times, allows for the early diagnosis of positive cases, preventing disease transmission ^(14)^. RATs may be administered by a healthcare worker or self-administered by the worker or patient. Self-administration, in which an individual collects a sample, runs the test, and interprets the results by themselves, is called “self-testing.”

In recent years, self-testing has gained prominence as an essential self-care intervention. In 2016, the World Health Organization (WHO) recommended HIV self-testing. Since then, self-testing for Hepatitis C (HCVST)(2021) and COVID-19(2022) has been recommended ^(16,17,18,19)^. Self-testing has proven to circumvent barriers to testing, such as stigma, privacy, time required for testing, and affordability ^(20, 21)^. Recent studies have demonstrated the accuracy of the COVID-19 self-test results ^(22, 23)^.

COVID-19 self-test products are approved for use in the general population in India but have not been widely used in workplace settings. In manufacturing industries, which face a shortage of healthcare workers on-site, self-testing may enable large-scale screening programs within workplaces, particularly among vulnerable workers, Such programs have the potential to improve worker health and mitigate pandemic-related business disruptions which can impact workers’ livelihood.

This study assessed the usability of a COVID-19 self-test in a peer-assisted self-testing model in two factories in Bengaluru, India. Peer assistance refers to a lay peer who was present during testing to guide workers through the testing procedure. This approach sought to balance the limitation of health worker availability with low literacy levels among the workforce. Additional acceptability assessments in the study determined how workers viewed the test and testing process.

## Materials and Methods

### Study Design

#### Location

This usability and acceptability study was a cross-sectional study conducted in Bengaluru, Karnataka, India, from 25^th^ January to 7^th^ April 2022 in two factories - 1) a mid-sized machine parts manufacturer with approximately 180 staff; and, 2) a leading garment manufacturer and export company with approximately 650 workers. Swasti, a global public health not-for-profit organization with existing workplace health programs in both factories, conducted this study. Factories were selected based on size, diversity of workforce demographics, and management’s willingness to implement a COVID-19 self-testing program designed to mitigate business disruptions.

#### Sampling

Factory management opened the testing event to their entire workforce. From those that presented at the testing event, the project coordinator recruited participants into the usability study if they met the inclusion criteria - all adults (over the age of 18 years) formally employed by the factories who provided informed consent. Senior Management and Healthcare Workers were excluded from the study due to variance in knowledge, education, and previous experience administering SARS-CoV-2 rapid antigen tests (Figure 1). Sampling was purposive to align with the underlying factory population. All participants were approached for a short quantitative survey after test completion to explore their individual preferences and experiences of peer-assisted self-testing. Four qualitative focus-group discussions were conducted with 17 factory workers who participated in the study. Participants were purposively sampled to ensure a diverse representation of age, sex, education, and roles (e.g., line workers, supervisors) in line with the sampling approach described for the study overall.

**Figure 1:**
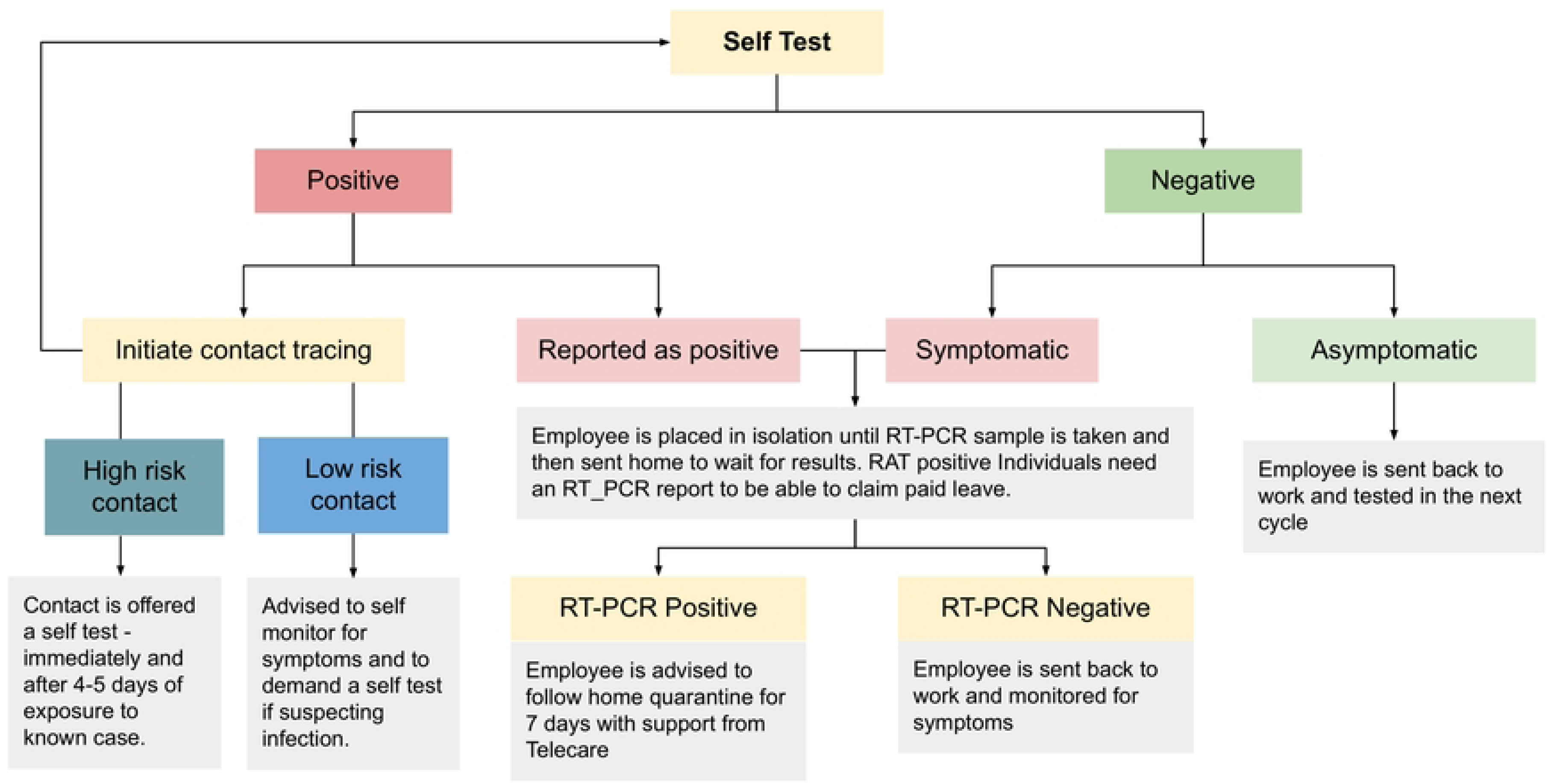
sample selection.

### Ethical Considerations

Ethical approval was received by the Catalyst Foundation Institutional Review Board on January 19, 2022. Participants were informed about the nature, purpose, and possible risks of the study and provided written informed consent.

### Study Procedures

Peers were selected based on specific criteria, including management recommendations, communication skills, learning ability or previous experience as peer educators, and literacy level to manage data entry support. Peers underwent training in infection control, proper use of personal protective equipment (PPE), correct administration of SARS-CoV-2 self-tests, and provision of support for test administration and result interpretation. They were also trained to provide a non-stigmatizing and supportive environment for self-testing. Additionally, peers learned how to use the manufacturer-provided app for reporting results and assisting participants in using the application.

Swasti coordinated the procurement and inventory of rapid antigen test kits for the usability study, ensuring proper temperature conditions during transit. The study used the Panbio™ COVID-19 Ag Rapid self-test kit, a nasal AgRDT COVID-19 Self-Test approved by the Indian Council of Medical Research (ICMR), the apex medical research organization that led and managed the COVID-19 response in India. Peers checked and ensured that only approved and valid test kits were used according to the manufacturer’s instructions. A detailed diagram of study procedures is provided (Figure 2).

**Figure 2:**
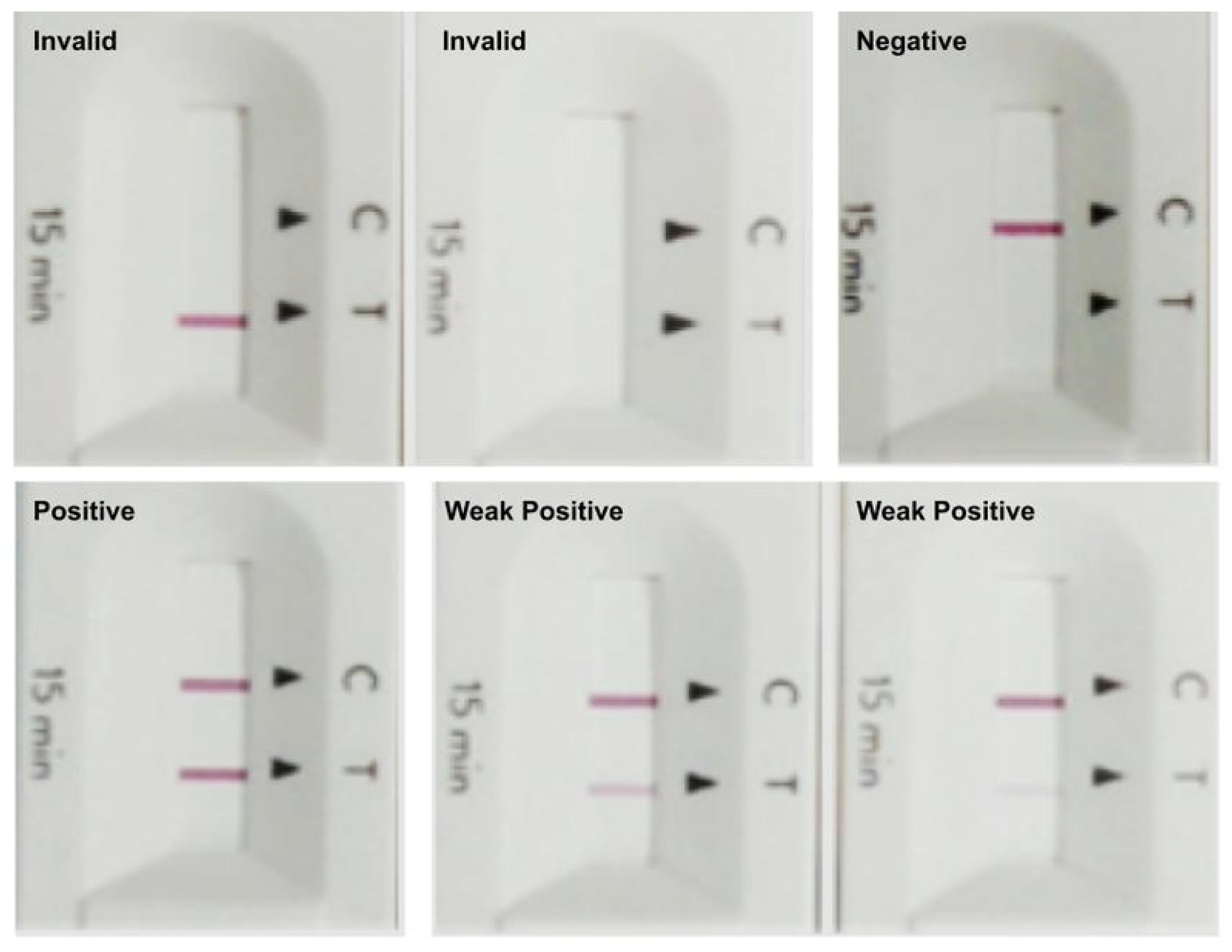
study procedures.

Trained peers and consented participants executed the following steps:

● The study commenced with a participant interview to capture demographic information.
● Participants were then given a self-test kit, personal protective equipment (a surgical mask, hand and surface sanitizer), and a data collection device with the NAVICA self-testing app for result reporting.
● The peer assistant provided an overview of the test components, orientation to the manufacturer-provided Instructions For Use (IFU), and guidance on result interpretation. They assisted the participants verbally as they performed each test step and offered demonstrations with their kit if the worker did not understand verbal instructions. The peers never touched the participant’s test during sample collection, test operation, or result interpretation
● During the 15-minute run time of the test, participants were asked to read a series of pictures representing strong positive, weak positive, negative, and invalid test results in a random order to assess result interpretation accuracy before they interpreted their test results.
● When the run time was complete, the observer, peer, and participant independently read and recorded the results.
● Participants were then asked to use the study devices to report their self-test results using Abbott’s NAVICA India^TM^ app. The app collects self-reported demographics, vaccination status, and COVID-19 exposure history and symptoms. Participants reported their test results using image capture. The image was auto-analyzed using artificial intelligence. The results were displayed and automatically shared with ICMR.
● No data was stored on the device, and neither the observer nor the peer assistant had access to the data reported via the app. Peer assistants helped participants report results via the NAVICA India^TM^ app, where required. All results were managed based on the national testing guidelines at the time (Figure 3). After completion of the test, the trained independent observers administered a quantitative survey to all the participants. A subset of the participants were invited to participate in FGDs to explore their reflections on self-testing.

**Figure 3:**
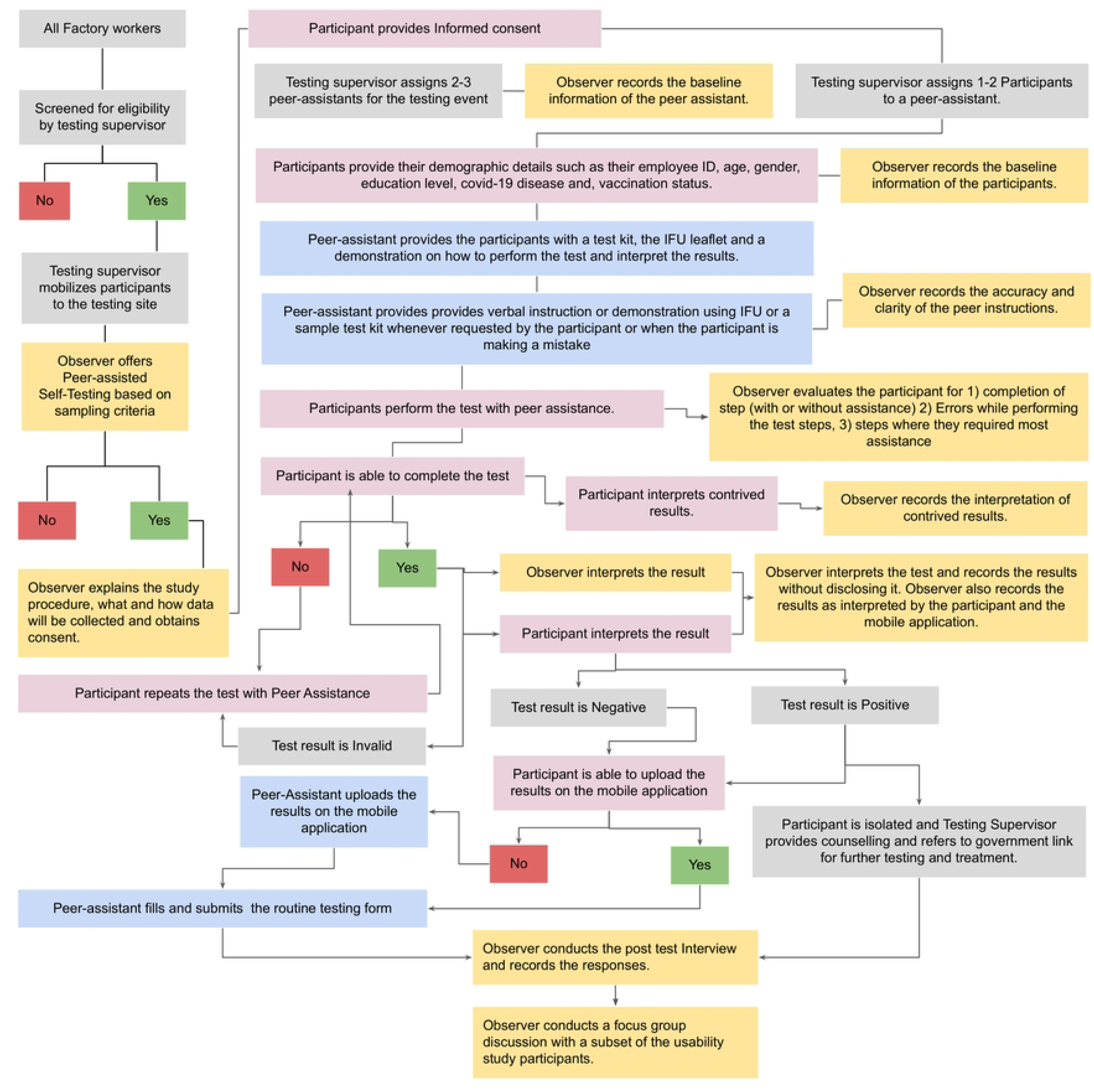
testing algorithm.

**Figure 4:**
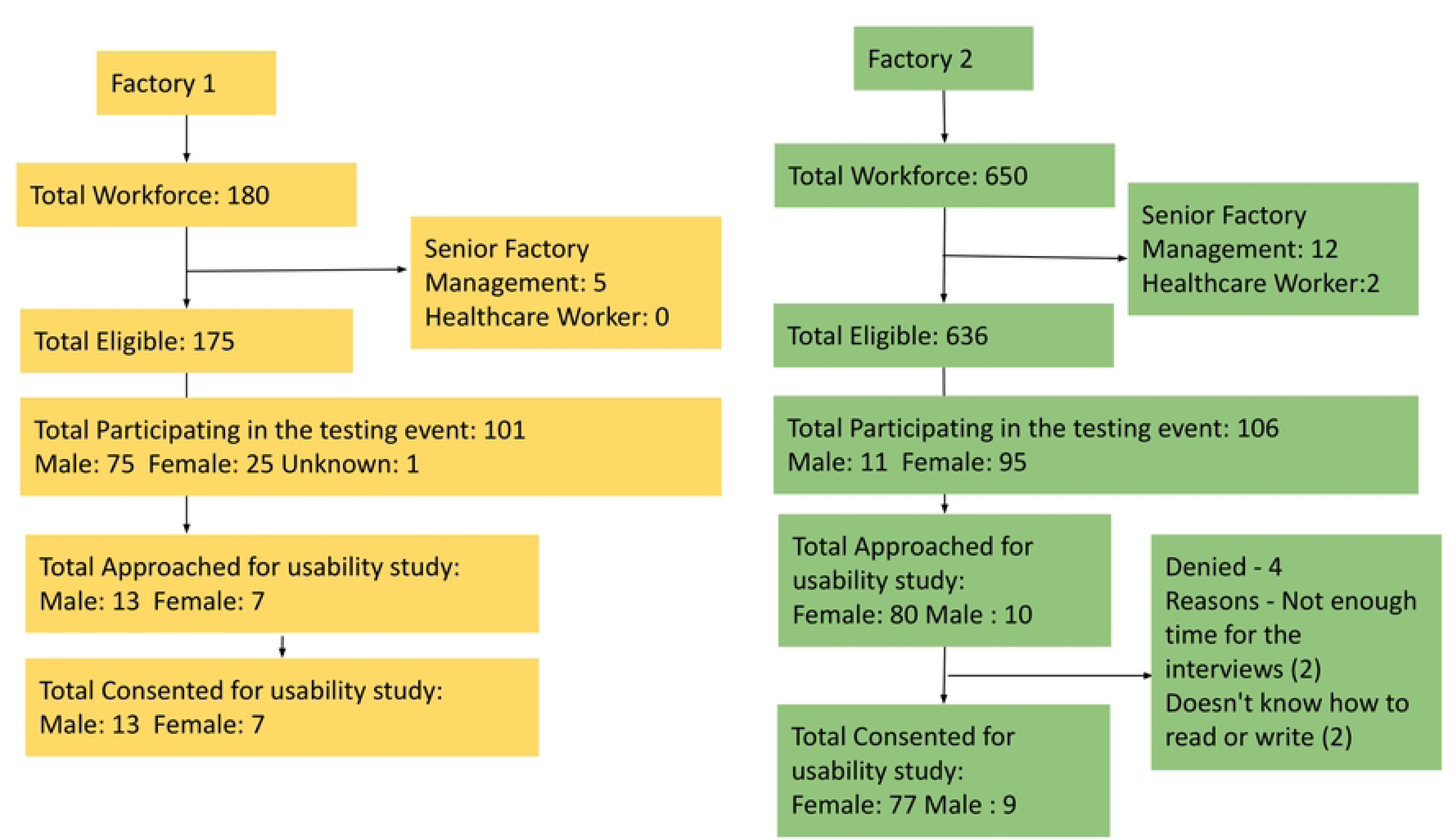
picture of contrived results.

### Data Collection

Highly trained and qualified researchers, known as observers, were deployed to evaluate the test operation by study participants and the support provided by peer assistants in both factories. These observers had expertise in conducting qualitative interviews and focus group discussions in the local language. They received comprehensive training on the study protocol, data collection tools, and the manufacturer-provided application. Observers employed a usability checklist guided by the Panbio Self-test IFU and adapted from tools used in other self-test usability studies <u>(Appendix 1)</u> ^(24)^. The checklist noted 13 critical and five non-critical steps. Observers used a separate section of the checklist to assess the quality of peer assistance provided. To assess correct interpretation, (1) The actual test result was read and recorded by both the worker and the observer, in addition to automated results read by the NAVICA app. The observer’s interpretation was considered the reference (2) The observer used the Result Interpretation section in the usability checklist <u>(Appendix 1)</u> to record whether the employee accurately interpreted the pictures of self-test results. Data collected through the paper-based usability checklist was entered into Microsoft Excel files, and the data entry quality was checked for accuracy and completeness.

Post-test Survey: After completing the test, all participants completed a short quantitative survey to explore their experiences and preferences for peer-assisted self-testing. They were also asked questions regarding post-test actions in case of both positive and negative results to determine whether they understood the instructions correctly. The observer also asked for comments about the study processes and the level of assistance required during and after the test. All data collection tools were tested internally for clarity and internal consistency.

A subset of study participants and peer assistants were invited to participate in focus group discussions (FGDs) in the days following the test. The observers conducted these discussions at the factory in a quiet and private training room, using a semi-structured guide and probes developed by the study team to explore specific themes of interest. Key areas explored included past experience with COVID-19 testing and preference for COVID-19 self-testing. The duration of the FGDs ranged from 20 - 30 minutes. All the audio transcripts were translated from the local language and transcribed into English through a third-party translator service.

### Data Analysis

#### Quantitative Analysis

Data were analyzed using IBM SPSS Statistics (v. 26.0). To analyze and describe participant and peer demographics, gender, age, and education were treated as categorical variables. To determine whether the samples from each factory resemble the overall demographics, statistical tests (t-test for age and chi-square test for gender and education levels) were performed. Sub-analyses by age, gender and factory were not performed due to the overall small sample size (N=106). Estimates of proportions of participants that completed the test steps correctly with peer assistance. (Usability index/ score) were calculated using the definitions and criteria described in the analysis plan <u>(Appendix 4)</u>. Usability scores for the test kit and the mobile application are reported separately. Estimates of the proportion of participants who correctly interpreted their actual COVID-19 self-testing results and pictures of contrived results are reported, as is the proportion of tests conducted with accurate peer support. The inter-reader agreement was also reported by the percentage of consistent results between each reader (participant, observer, and mobile application). All questions, responses, and corresponding frequencies are listed in <u>(Appendix 5).</u>

#### Qualitative Analysis

We explored the qualitative data using thematic and discourse analysis approaches. Two researchers first read the transcripts to understand the participants’ views and preferences, considering the social context in which the discussions were conducted. In the second stage, the researchers identified concepts or codes inductively from the data. After one round of dialogue between both researchers, a set of codes were agreed upon and defined. In the next stage, relationships between the codes were described, and larger categories of codes were formed. The generated code categories and the attached definitions and text excerpts were reviewed with two other coders (Senior Researchers) to come to a consensus about the themes generated and modify any coding discrepancies through one more round of deductive coding. The latest version of NVivo 1.6.1 (1137) was used for analysis.

## Results

### Demographics

A total of 106 individuals consented to participate in the study. The overall mean age of the sample was 35.7 yrs. More women than men participated in the study (Female: 84 (79.25%), Male: 22 (20.75%)). Just over half the sample had finished High school (12^th^ grade) (52.9%) <u>(Table 1)</u>. Demographics within each factory sample (Factory 1: N=20 and Factory 2 N=86) corresponded with the overall factory demographics <u>(Table 2)</u>. All of the participants had yet to use a COVID-19 self-test before. Only 7(6.6%) participants noted that they had used a self-test for indications other than COVID-19 before (e.g. Pregnancy test).

**Table 1:**
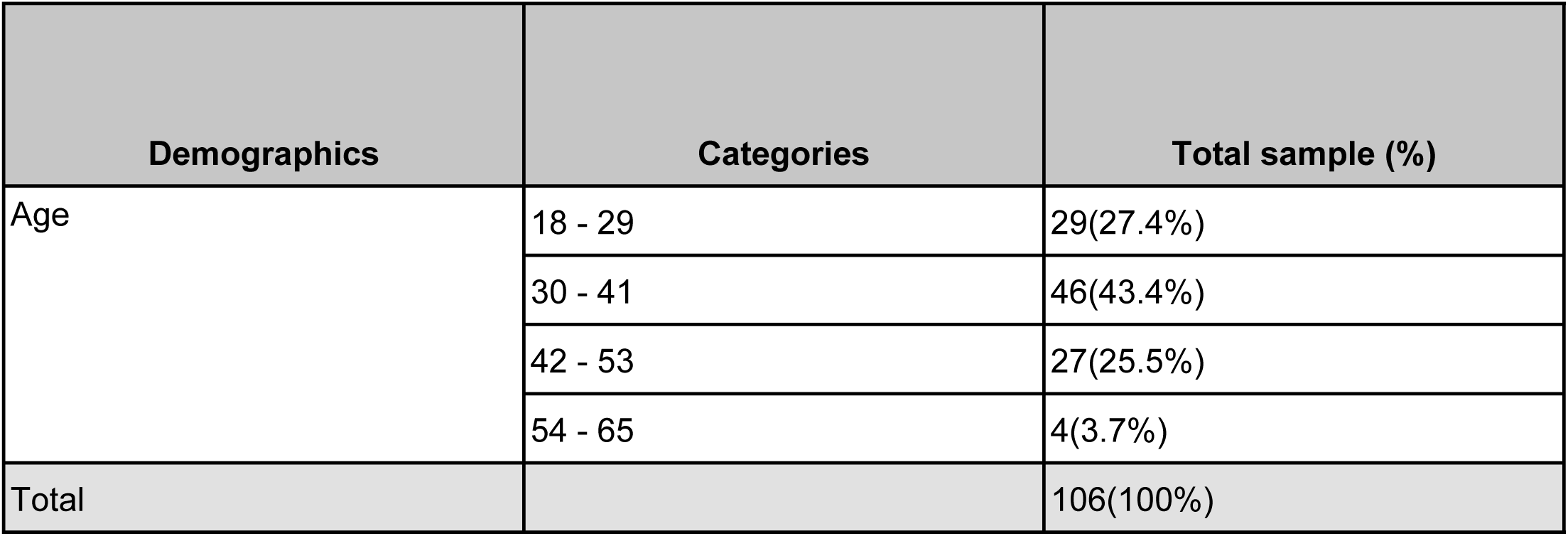
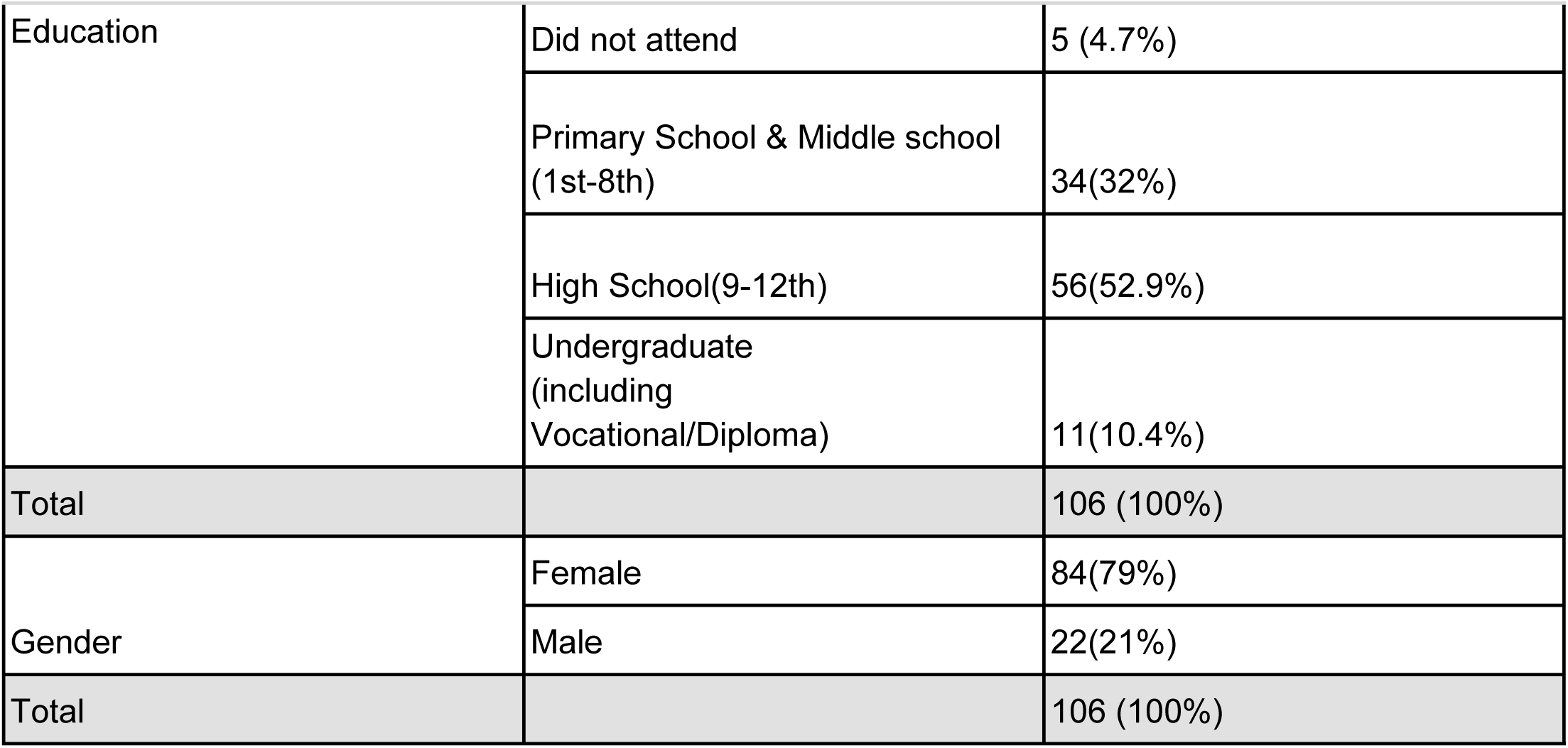
Participant Demographics.

| Demographics | Categories | Total sample (%) |
| --- | --- | --- |
| Age | 18 - 29 | 29(27.4%) |
|  | 30 - 41 | 46(43.4%) |
|  | 42 - 53 | 27(25.5%) |
|  | 54 - 65 | 4(3.7%) |
| Total |  | 106(100%) |
| Education | Did not attend | 5 (4.7%) |
|  | Primary School & Middle school (1st-8th) | 34(32%) |
|  | High School(9-12th) | 56(52.9%) |
|  | Undergraduate (including Vocational/Diploma) | 11(10.4%) |
| Total |  | 106 (100%) |
| Gender | Female | 84(79%) |
|  | Male | 22(21%) |
| Total |  | 106 (100%) |

**Table 2.**
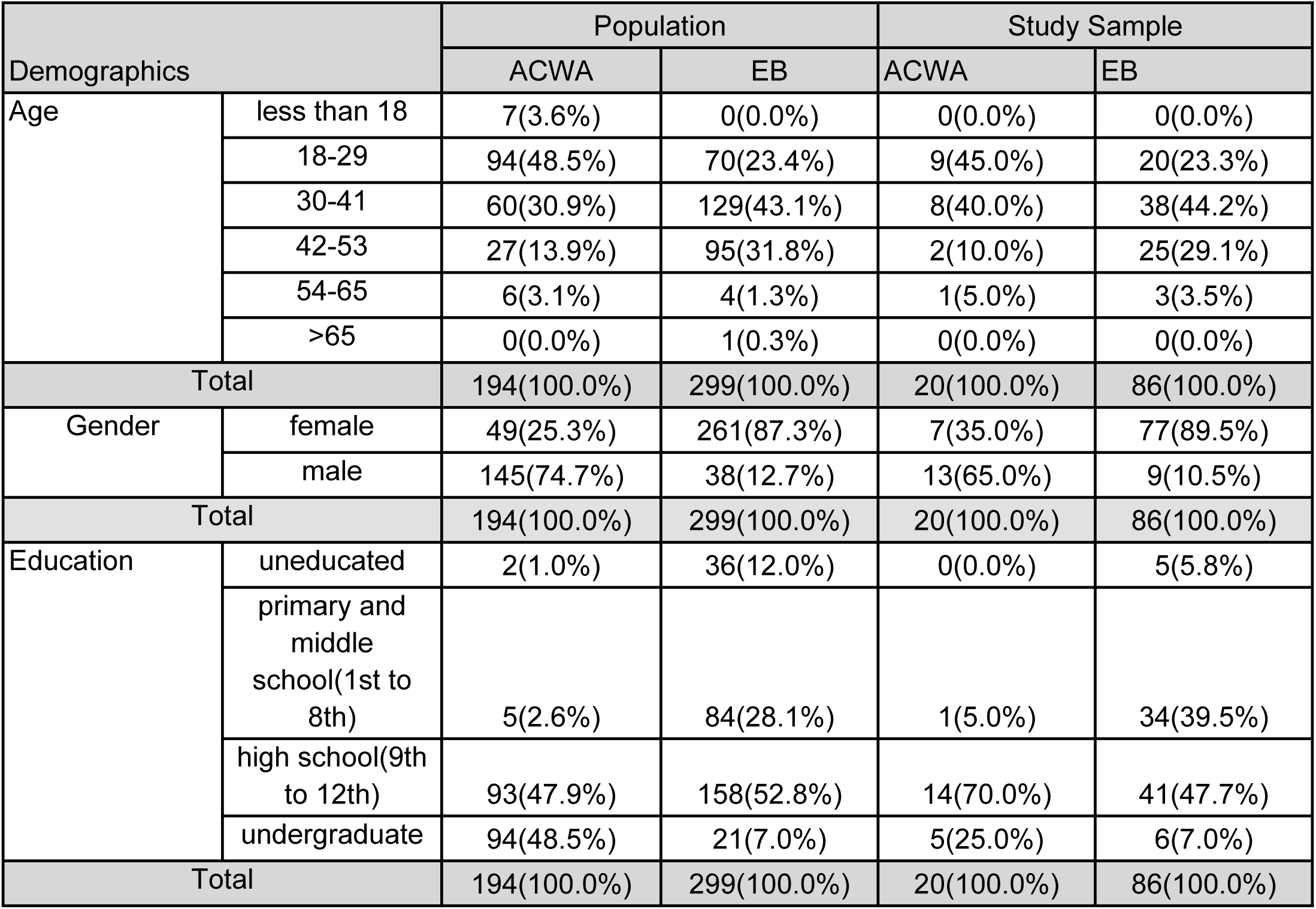
Population and sample demographics.

| Demographics |  | Population |  | Study Sample |  |
| --- | --- | --- | --- | --- | --- |
|  |  | ACWA | EB | ACWA | EB |
| Age | less than 18 | 7(3.6%) | 0(0.0%) | 0(0.0%) | 0(0.0%) |
|  | 18-29 | 94(48.5%) | 70(23.4%) | 9(45.0%) | 20(23.3%) |
|  | 30-41 | 60(30.9%) | 129(43.1%) | 8(40.0%) | 38(44.2%) |
|  | 42-53 | 27(13.9%) | 95(31.8%) | 2(10.0%) | 25(29.1%) |
|  | 54-65 | 6(3.1%) | 4(1.3%) | 1(5.0%) | 3(3.5%) |
|  | >65 | 0(0.0%) | 1(0.3%) | 0(0.0%) | 0(0.0%) |
| Total |  | 194(100.0%) | 299(100.0%) | 20(100.0%) | 86(100.0%) |
| Gender | female | 49(25.3%) | 261(87.3%) | 7(35.0%) | 77(89.5%) |
|  | male | 145(74.7%) | 38(12.7%) | 13(65.0%) | 9(10.5%) |
| Total |  | 194(100.0%) | 299(100.0%) | 20(100.0%) | 86(100.0%) |
| Education | uneducated | 2(1.0%) | 36(12.0%) | 0(0.0%) | 5(5.8%) |
|  | primary and middle school(1st to 8th) | 5(2.6%) | 84(28.1%) | 1(5.0%) | 34(39.5%) |
|  | high school(9th to 12th) | 93(47.9%) | 158(52.8%) | 14(70.0%) | 41(47.7%) |
|  | undergraduate | 94(48.5%) | 21(7.0%) | 5(25.0%) | 6(7.0%) |
| Total |  | 194(100.0%) | 299(100.0%) | 20(100.0%) | 86(100.0%) |

### Analysis of Key Outcomes

#### Usability of the Self-Test

##### Usability of the SARS-CoV-2 Self-Test in a Peer-Assisted Model

The usability of the self-test was 75.9% for the overall sample (N=103) across all steps and 80.7% (N=103) when restricted to critical steps <u>(Table 3)</u>. The observer checklist data was not available for three individuals. The most commonly reported errors (<75% participants performed accurately) were:

1. Using the incorrect amount of buffer
2. Inaccurate swab insertion
3. Failure to swab the nose five times, touching the walls.

**Table 3.** Usability Index - Test Kit.

| Steps | The proportion of participants that performed a step correctly (N = 103) |  |  |
| --- | --- | --- | --- |
|  | n | Yes | No |
|  |  | Total | Total |
| Was the employee able to squeeze the liquid from the Buffer Bottle into the Tube? | 97 | 84(86.6%) | 13(13.4%) |
| Did the participant confirm that the buffer level was above the line provided in the tube? | 97 | 70(72.16%) | 27(27.84%) |
| Was the employee able to remove the swab without touching the soft end? | 98 | 89(90.82%) | 9 (9.18%) |
| Did the employee insert the soft end up to 2 cm into their nostrils? | 98 | 57(58.16%) | 41(41.84%) |
| Did the employee rotate the swab five times in each nostril? | 97 | 69(71.13%) | 28(28.87%) |
| Did the employee insert the swab into the fluid and swirl it five times while pushing against the tube wall? | 98 | 74(75.51%) | 24(24.49%) |
| Was the employee able to place five drops (and not more or less) into the well? | 96 | 79(82.29%) | 17(17.71%) |
| Did the employee place the solution into the well on the test device (and not any other location)? | 96 | 89(92.71%) | 7 (7.29%) |
| Did the employee read the results within 15-20 minutes? | 95 | 92(96.84%) | 3 (3.16%) |
| **Usability Index total** |  | **80.7%** |  |

##### Usability of the App

The usability of the NAVICA mobile application to report the test results was 34.0% (N=103) <u>(Table 4)</u>. Less than 30% of participants scored accurately in the following steps:

1. Enter all the mandatory data fields in the mobile application to proceed with result capture.
2. Capture the results for automatic upload into the mobile app

**Table 4.** Usability Index - Mobile Application.

| Steps | Proportion of participants that performed a step correctly (N = 103) |  |  |
| --- | --- | --- | --- |
|  | n* (no of valid observations) | Yes | No |
|  |  | Total | Total |
| Did the employee successfully sign up for the application? | 95 | 30(31.6%) | 65(68.4%) |
| Could the employee enter all the details required to move to the next steps? | 95 | 27(28.4%) | 68(71.6%) |
| Could the employee read the results analysed by the application? | 92 | 47(51.1%) | 45(48.9%) |
| Did the employee finish uploading the results on the application? | 92 | 22(23.9%) | 70(76.1%) |
| **Usability Index (Weighted Average of the usability scores of steps)** |  | **33.80%** |  |

##### Reported ease of use

Fifty-three study participants (52.5%) (N=101) in both factories reported that the test was easy to conduct. Thirty-seven (36.6%) (N=96) participants said it was somewhat easy, and 11(10.9%) participants said it was not accessible at all (<u>Table 5</u>). Usage of the mobile application was the step that most participants reported needing assistance with (45.4%), followed by sample collection (21.6%) and preparing the buffer and test kit elements (20.6%) (<u>Table 5</u>). This corresponds with the steps where most participants performed errors.

**Table 5:** Reported ease of use and assistance required.

|  |  | Total |
| --- | --- | --- |
| How easy was the test to conduct? | a. Not easy at all | 11(10.9%) |
|  | b. Somewhat easy | 37(36.6%) |
|  | c. Completely easy | 53(52.5%) |
| Total |  | 101(100.0%) |
| Which parts of the test did you find difficult and require assistance with? | a. Test Preparation (preparing buffer and test kit elements) | 20(20.8%) |
|  | b. Sample Collection (collecting sample through nasal swab) | 17(17.7%) |
|  | c. Test Process (placing buffer with sample in well) | 13(13.5%) |
|  | d. Use of Mobile Application | 40(41.7%) |
|  | e. Result Interpretation | 6(6.3%) |
| Total |  | 96(100.0%) |

#### Interpretation of the Test

##### Inter-reader concordance

<u>Table 6</u> depicts the percentage of concordance in test result interpretation between the manufacturer’s mobile application, participants, and observers (N=95). Five participants deferred to the peer assistant/observer to interpret the results, and the app did not analyze six results due to a technical error and were not included in this sample. In all instances of discordance, the observer’s interpretation was considered final.

1. Concordance between participant and observer was 97.9% (93). The two instances of discordance were due to an incorrect interpretation of weak positive results by the participant.
2. Concordance between the participant and the application was 97.9% (93). The two instances of discordance were due to inaccurate interpretation by the mobile application in which negative results were read as positive results.
3. Concordance between the observer and the application was 95.8% (91). 50% (n=2) of discordance was due to the app’s interpretation of a negative test result as a positive one. 50% of discordance was due to the app’s interpretation of a positive test result as a negative one.

**Table 6.** Inter-Reader Concordance for interpretation of results.

| Inter-Reader Concordance for interpretation of results | % of concordance | % of discordance |
| --- | --- | --- |
| Participant interpretation vs observer interpretation | 93(97.9%) | 2(2.1%) |
| Participant interpretation vs manufacturer app interpretation | 93(97.9%) | 2(2.1%) |
| Observer interpretation and manufacturer app interpretation | 91(95.8%) | 4(4.2%) |

##### Interpretation of Contrived Results

Most participants accurately interpreted the pictures of strong positive and negative results (∼82.9% and ∼80%, respectively). 69 (65.7%) participants interpreted weak positive results correctly, whereas 70 (66.7%) interpreted invalid results correctly. The proportions of participants who did and did not interpret the four types of potential results are detailed in <u>Table 7</u>.

**Table 7.** The proportion of participants who correctly interpreted SARS-CoV-2 self-testing results using high-resolution pictures of self-test results by gender.

| Self-test results | Proportion of participants who interpreted the results correctly | Proportion of participants who did not interpret the results correctly |
| --- | --- | --- |
| Negative | 84(80.0%) | 21(20.0%) |
| Weak Positive | 69(65.7%) | 36(34.3%) |
| Positive | 87(82.9%) | 18(17.1%) |
| Invalid | 70(66.7%) | 35(33.3%) |

#### Peer Support Model

The study involved ten peers who assisted participants during self-testing. Their mean age was 33.2 years. They were predominantly female (60%). More than half of the peers (70%) had completed high school (grades 9-12), peer assistant demographics are detailed in <u>Table 8</u> below.

**Table 8.**
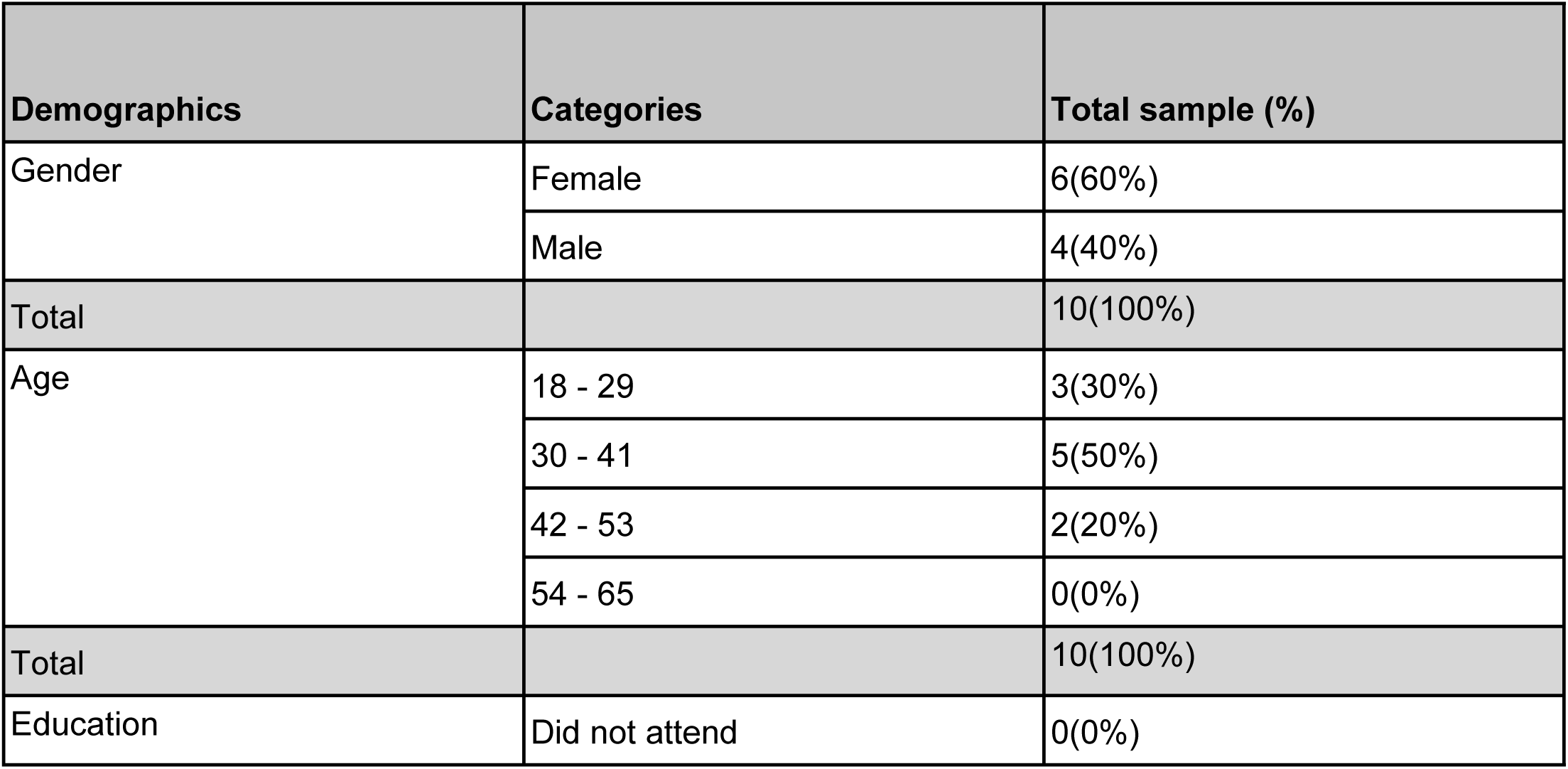
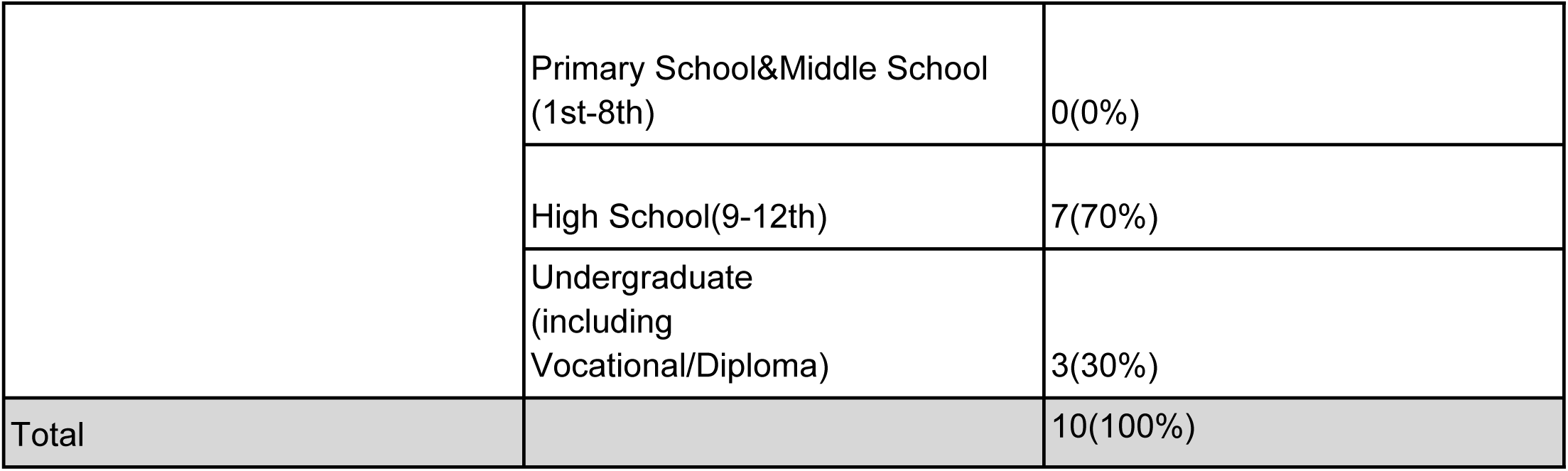
Peer assistant Demographics - N-10.

##### Peer Assistance Efficiency

Across all critical steps, peer assistants provided accurate instructions in 93.4% of tests performed. The number and proportion of tests for which the peer assistant performed an error are reported for each critical step in <u>Table 9</u>. Among the steps critical to the test, peers missed or provided incomplete instructions for sample collection and verifying appropriate buffer levels. A list of the most common errors reported in peer assistance is in <u>Table 10</u>. 53.9% (55) of participants from both factories found the verbal peer instructions *“Completely easy”* to understand, 36(35.3%) found the instructions *“Somewhat easy”, and* 11(10.8%) found them *“Not easy at all”*

**Table 9.** Peer Assistant Efficiency.

| Critical Steps | Proportion of tests for which the peer provided accurate support (N=103) |  |  |
| --- | --- | --- | --- |
|  | n (no of valid observations) | Yes | No |
| Did the PA* carefully explain different components of the test kit (swab, buffer bottle, tube, test device, tube rack, disposable bag, timing device). | 99 | 91(91.9%) | 8(8.1%) |
| Did the PA instruct the employee to empty the buffer into the tube to avoid spillage? | 98 | 98(100.0%) | 0(0.0%) |
| Did the PA check if the buffer is above the line marked in the tube? | 98 | 80(81.6%) | 18(18.4%) |
| Did the PA provide instructions to unwrap the sample collection swab appropriately? | 98 | 96(98.0%) | 2(2.0%) |
| Did the PA provide instructions to use the sample collection swab appropriately? | 98 | 96(98.0%) | 2(2.0%) |
| Did the PA instruct you not to touch the swab at any point? | 98 | 87(88.8%) | 11(11.2%) |
| Did the PA check with the employee to ensure they inserted the swab to the correct depth? | 97 | 91(93.8%) | 6(6.2%) |
| Did the PA check with the employee to ensure they rotated the swab five times in each nostril? | 97 | 95(97.9%) | 2(2.1%) |
| Did the PA check with the employee to ensure they inverted the swab into the buffer tube without putting it down anywhere? | 97 | 95(97.9%) | 2(2.1%) |
| Did the PA instruct the employee to check the liquid for bubbles? | 98 | 64(65.3%) | 34(34.7%) |
| Did the PA instruct the employee to place five drops (and not more or less) into the well and not any other location on the test? | 94 | 93(98.9%) | 1(1.1%) |
| Did the PA alert the employee to read their results in the accurate read time window? | 95 | 94(98.9%) | 1(1.1%) |
| Did the PA instruct the employee how to interpret their results? | 94 | 94(100.0%) | 0(0.0%) |
| Did the PA give instructions about reporting results? | 96 | 94(97.9%) | 2(2.1%) |
| **Average score** |  | **93.4%** |  |

\*PA= peer assistant

**Table 10:** Errors reported in Peer Assistance.

|  |  |
| --- | --- |
| Common errors observed via the checklist:<br>• Peer did not provide instructions to wear a mask correctly at all times except sampling<br>• Peer did not provide instructions to check if an accurate amount of buffer is being used<br>• Peer did not provide instruction not to touch the swab at any point<br>• Peer did not provide instructions to check liquid for bubbles | Qualitative data and informal observation suggest additional errors:<br>• Peer does not speak loudly<br>• Peer touches the participant's test kit to demonstrate a step<br>• Peer is giving instructions too fast<br>• Peer is giving incomplete instructions<br>• Peer does not explain the context or meaning of certain instructions |

#### Acceptability of the Test

##### Confidence in using the test kit

**With peer assistance:** 62% (62) of participants reported they were completely confident in using the test and interpreting self-test results with the assistance of a peer. 28.0% (28) were somewhat confident and 10.0% (10) participants were not confident at all.

**Unassisted:** 43.9% (43) participants were completely confident in performing and interpreting the test on their own, 46.9% (46) participants were somewhat confident, and 9.2%(9) participants were not confident at all.

##### Confidence in using the mobile application

**With peer assistance:** Only 15.6% (15) participants were completely confident in capturing and uploading their results using the NAVICA mobile application with peer assistance. In comparison, 29.2% (28) and 55.2% (53) participants said they were somewhat confident and not confident at all, respectively.

**Unassisted:** Only 19.6% (19) participants were completely confident in capturing and reporting their results independently. 35.1% (34) participants and 45.4%(44) participants stated they were not confident in capturing and uploading their results using the mobile application without assistance.

##### Accessing the Test

57.4% (58) participants reported that they would prefer to use the self-test at home, followed by 36.6% (37) who preferred to test at the workplace. Finally, 5.0% (5) of participants preferred to test at a hospital. When asked where participants wished to purchase a test, 36.1% (35) of participants preferred their nearest pharmacy. 29.9%(29) participants preferred to access the test at the workplace, and 14.4%(14) participants would like to access the test at the PHC/local clinic. All the questions in the post-test interview, responses, and frequencies are linked in <u>(Appendix 5).</u>

#### Qualitative Results

Motivations and barriers to self-testing, perceived benefits of self-testing, and reflections on the peer-assisted self-testing model were the key themes that emerged from the qualitative thematic analysis of focus group discussions with the participants.

##### 1. Motivators and barriers for Self-testing

All participants were first-time users of a COVID-19 self-test. Participants reported being fearful of testing but were motivated to accept and perform self-testing due to concerns about getting infected and spreading the infection to others. In both factories, participants’ hesitancy to test was precipitated by the fear of a positive result resulting in isolation from their peers and the stigma associated with working with an infected person.

> *"When we are in close contact with infected individuals, there is a risk of transmission to us. This is why we should undergo testing." "It is important to prioritize testing to prevent harm or danger to others." (Male, 25 years, Factory 1).*

> *“If any of us test positive, it may instill fear among others, leading to hesitation in working together.” (Female, 21 years, Factory 2)*

##### 2. Perceived Benefits of Self-testing

Participants in both factories showed mixed sentiments about the COVID-19 self-testing experience, with generally positive attitudes toward the test. Several participants spoke about their experiences with other testing methods (RT-PCR and professionally administered RAT) and compared them with their self-testing experience. Specifically, participants preferred collecting samples from the nose (as opposed to nasopharyngeal collection), describing it as a comfortable and painless procedure.

> *Sometimes the testing will be painful. But, if we do it on our own, it is not painful. So we feel this is better. (Female, 32 years, Factory 1)*

Most respondents mentioned that self-testing saved time and reduced risks posed by the long wait for testing in government facilities. Participants thought that self-testing at the workplace was quick and easy in terms of time taken to set up, test and interpret results.

> *If we go to a Bruhat Bengaluru Mahanagara Palike (BBMP - Local administrative body in the Greater Bengaluru metropolitan area) test center, we are unsure of the safety. There were long queues, and we doubted whether someone among these people was infected. (Male, 25 years, Factory 1).*

> *One point is that we can do it easily and leisurely at work. When we go somewhere else, it takes too much time. (Female, 32 years, Factory 1)*

Participants highlighted that the ability to understand the process and to do the test at home with family members is one of the critical benefits of the self-testing kit.

> *“We can understand this (self-testing process). We can do this at our houses when our husbands or children or relatives are infected. Or we can guide them on how to do the testing.” (Female, 35 years, Factory 2)*

##### 3. Reflections on Peer-Assisted Self-Testing Model

Participants also shared their experiences with the specific peer-assistance models used during self-testing in factories. Participants from both factories claimed that peer assistance was a critical component of self-testing, finding comfort in performing the test after receiving in-person instructions from their peers. Having colleagues as peer instructors was preferred over healthcare workers, as instructions provided by their co-workers were straightforward to understand and follow.

> *“When we went to a government hospital, they simply took the swab and sent us back. They did not explain anything. Here (at the workplace), they (peers) have explained it to us.” (Female, 35 years, Factory 2)*

Some participants mentioned that peer assistants should have more experience and knowledge about the tests, most likely someone with healthcare experience. Others highlighted that their co-workers would need more training and handholding support from the study staff to perfect their support.

> *“We prefer your team (Swasti team) as you have more knowledge than our co-workers. They (peer assistants) have been trained very recently, and their knowledge is quite different. It is better if we learn something from you.” (Male, 21 years, Factory 1)*

> *We were comfortable as our co-workers in the company were providing us with instructors. (Female, 24 years, Factory 1)*

In addition to peer support, participants reported that more time to perform and interpret the results would also strengthen testing quality.

> *“If we are taking time from work, our attention will be split between work and testing, and we might make mistakes. But if we have good enough time, we will do it better and with concentration.” (Female, 24 years, Factory 1)*

## Discussion

This study assessed the usability of antigen-based SARS-CoV-2 self-testing in a peer-assisted model among factory workers - a key vulnerable group facing inequitable access to early diagnosis and testing. Prior research on self-testing for SARS-CoV-2 evaluated unassisted and healthcare worker-assisted models in various settings, including educational institutions (25) (26), hospitals (27), and the general population (28). The majority of these studies were limited to high-income settings. While workplace models have been described, these models have relied on healthcare worker administration, home-based self-testing, or secondary distribution to household members (29, 30, 31). This is the first study to describe peer-assisted self-testing in a workplace setting where increased user support may be required due to reduced literacy levels.

Our study found that most factory workers could accurately conduct the critical steps for a nasal sampling-based test kit (80.7%) and interpret their results accurately with high levels of concordance (93.7%) with trained observer-interpreted results. This is comparable to the work by Sibanda et al. (23), who found the usability of the Panbio self-test on 12 critical steps to be 90.4% and 70.6% in Malawi and Zimbabwe, respectively. In our study, most participants reported the test was easy to conduct but required assistance in results interpretation, followed by sample collection. In line with usability data from Lesotho and Zambia, the common errors included failure to insert the swab to the correct depth, the inadequate swirling of the swab to touch the nasal passage walls, and buffer preparation (32). Low usability scores on some of the critical test steps in our study could have been driven by multiple factors such as demographics, first-time use of a self-test, clarity of peer instruction, nature of sample collection, and limited time for testing due to the workplace setting (33, 34).

Simple changes to test design could address some usability challenges. The design for SARS-CoV-2 self-test kits did not include populations with limited education and English-speaking abilities. Instructions need to be printed in local languages. Using a quick reference guide could target key usability challenges and improve the accuracy of SARS-CoV-2 test interpretation (35). Instructional videos and pictures have also been suggested to improve the usability of other self-tests. (34) (36). Providing a pre-filled extraction tube or designing an easy-to-open cap for the buffer bottle and enhancing the clarity of the buffer line could also address some user errors. Such work will be critical to bring the reported usability of SARS-CoV-2 self-tests in-line with other self-test products, such as oral and blood-based HIV self-tests (37).

Reported challenges with the instructions for use underscore the importance of peer assistance for users who reported reduced confidence in performing the test without peer assistance. Data on the accuracy of peer instructions suggests that workplace testing programs must closely and continuously monitor the quality of peer instruction and focus on ensuring strong self-efficacy among peers via regular refresher training with online/self-paced modules, toolkits, job aids, and supervision by management. These interventions can strengthen accurate use and decrease the time required for testing services, a critical priority reported by management. Such investments will strengthen the role of peers, who are essential in driving the uptake of self-testing services in the workplace (38; 39). As these tests are used more frequently, individuals are expected to gain more confidence in using them independently without relying on peer support, especially in places they prefer, such as their homes. Similar results have been documented in workplace settings in Kedah, Malaysia (31).

This study demonstrated that time constraints can influence the peer’s quality of instruction and the participant’s focus on the test, resulting in instructional and testing errors. Workplace programs, therefore, need to ensure that workers have sufficient testing time and privacy to interpret results in tandem with withdrawing workers from an active production line.

We found strong concordance in test interpretation between users and trained observers (97.9%). Discordance between users and observers was driven by incorrect user interpretation of weak positive results. Low accuracy in the interpretation of contrived results underscores poor understanding of weak positive and invalid results. Result interpretation errors, especially in contrived image review, were most likely due to an insufficient wait time for interpretation, not having the IFU in a local language for reference, and lack of attention during peer support. These findings align with the broader self-test literature, including tests for SARS-CoV-2, underscoring the potential for misinterpretation of these test outcomes (40, 41).

Concordance between the mobile application and trained observers was lower (95.8%) and driven by false interpretations by the app. In the case of false negatives, participants were often hesitant to accept the interpretation of the trained observer and required additional counseling or re-testing. We hypothesize that false results may have been driven by improper lighting or poor focus for image capture. This aligns with our findings that the usability of the manufacturer-provided (NAVICA^TM^) app for results reporting in a factory setting is low. Participants and the peer assistants faced multiple challenges with an app-based model for reporting, including unavailability of content in the local language, multiple mandatory fields, usage of technical and hard-to-understand words, and frequent technical errors. Findings suggest that a mobile-based reporting model has low usability and is moderately accepted by factory workers and similar populations with restricted smartphone access and digital literacy. Industrial workers cannot carry their mobile phones to their workspace, limiting their ability to report and view results on their devices. Our findings are similar to recent research [17] [18] that suggests mobile application-supported HIVST is feasible only for youth and tech-savvy populations (42) (43). Point-of-care diagnostic manufacturers must be cognizant of these challenges if product penetration reaches the last mile and is acceptable to vulnerable populations. Self-test program implementers must design programs to circumvent these limitations, reducing dependencies on worker devices by providing program devices, stable internet, and peer support to mitigate technical errors and minimize false results.

Our findings also demonstrate that SARS-CoV-2 continues to offer end users many of the same benefits of self-testing for other infectious diseases, including knowledge of health status, increased comfort, and reduced time requirements (31, 44). These data add to a growing body of evidence demonstrating user preference for self-care technologies (45).

With health worker shortages and a lack of access to affordable healthcare services, more significant investments are needed in making community-centric self-care tools, including self-tests, more user-friendly, affordable, and acceptable. These findings indicate that self-testing is a feasible tool to facilitate manufacturer implementation of worker screening programs. It also allows vulnerable groups like factory workers to take charge of their health by monitoring their well-being, making informed healthcare choices, and improving communication and advocacy with employers and healthcare providers. Self-test technology should be preparative for future pandemics and support health system resilience. By reaching hard-to-reach groups, the peer-assisted self-testing model may provide an approach to expanding access to address the country’s unique needs.

## Limitations

To minimize participant selection bias and allow generalizability, sampling criteria were clearly defined to match the demographics of the target population in factories. The study could not assess the acceptability of reporting as reporting was mandatory. The presence of research staff could induce performance stress leading to mistakes by both the peer assistants and workers. To mitigate this, observers were instructed to be discreet and not interfere in the procedure. Standardized training followed by assessing their proficiency was provided to all peers to minimize the impact of varying quality of peer assistance on usability. To minimize the interpretation bias that occurs due to having multiple readers (observer, participant, peer assistant, and the mobile application), the participants’ interpretation was recorded before having the app or observer read and record the result. The observer did not disclose the results that s/he interpreted to the participant to prevent interpretation bias further and avoid any impact on acceptability responses. The small sample size limited our ability to assess the impact of demographics on usability and limited the generalizability of qualitative data on acceptability.

## Conclusion

These data add to a growing body of research demonstrating the usability, feasibility, acceptability, and cost-effectiveness of workplace testing, including self-testing, for SARS-CoV-2 (29, 30, 31, 46). They are the first to demonstrate the usability of SARS-CoV-2 self-tests in a peer-assisted model in factory settings with disadvantaged populations. Our findings suggest that peer-based models ensure that SARS-CoV-2 self-tests are usable with strong indications of acceptability. Determining the feasibility of implementing this model at scale is the next step to inform and advocate for other workplace self-testing programs for vulnerable populations.

## Data Availability

The quanitative data underlying this papers findings will be made available upon request to the corresponding author. FIND does not share qualitative data due to our qualitative data sharing standards. FIND is developing a quanitative data sharing policy and once that policy is developed and approved then FIND will be able to share data to open repositories in line with said data sharing policy

## List of Abbreviations

COVID-19: Coronavirus disease 2019
SARS-CoV-2: Severe Acute Respiratory Syndrome CoronaVirus-2
GDP: Gross Domestic Product
RT-PCR: Reverse transcription polymerase chain reaction
RAT: Rapid Antigen Test
WHO: World Health Organization
ICMR: Indian Council of Medical Research
GOI: Government of India
PPE: Personal Protective Equipment
AgRDT: Antigen based Rapid Diagnostic Test
PHC: Primary Health Center
BBMP: Bruhat Bengaluru Mahanagara Palike
IFU: Instructions for Use
CHW: Community Health Worker

## Author Contributions

Meghana Ratna Pydi: Data Curation, Formal Analysis, Investigation, Project Administration, Visualization, Writing – Original Draft Preparation, Writing – Review & Editing. Petra Stankard: Conceptualization, Writing – Original Draft Preparation, Writing –Review & Editing, Methodology, Validation, Visualization. Neha Parikh: Project Administration, Supervision, Validation, Writing – Original Draft Preparation, Writing – Review & Editing. Purnima Ranawat:-Conceptualization, Project Administration, Supervision, Writing – Review & Editing. Ravneet Kaur: Formal Analysis, Writing – Review & Editing. Shankar AG: Project Administration, Investigation, Writing – Review & Editing. Angela Chaudhuri: Supervision, Funding Acquisition, Writing – Review & Editing. Sonjelle Shilton: Conceptualization, Writing – Review & Editing, Methodology, Validation. Aditi Srinivasan: Project Administration, Funding Acquisition, Writing – Review & Editing. Joyita Chowdhury: Project Administration, Writing – Review & Editing. Elena Ivanova Reipold: Conceptualization, Writing – Review & Editing, Methodology, Validation.

## REFERENCES

1. WHO. WHO Health Emergency Dashboard. In: WHO, editor. Daily. Geneva2020.

2. Nexidigm private limited. (2021, June 30). Impact Of COVID-19 On The Manufacturing Sector In India. Mondaq. https://www.mondaq.com/india/operational-impacts-and-strategy/1086172/impact-of-COVID-19-on-the-manufacturing-sector-in-india-

3. ILO, Work B. Gendered impacts of COVID-19 on the garment sector. Geneva: International Labour Organization; 2020.

4. Raju E, Dutta A, Ayeb-Karlsson S. COVID-19 in India: Who are we leaving behind? Progress in Disaster Science. 2021;10:100163.

5. 754 factories shut, 46,000 lost jobs in 2 years in Karnataka. (2022, Mar 30). The times of India. https://timesofindia.indiatimes.com/city/bengaluru/754-factories-shut-46000-lost-jobs-in-2-years-in-karnataka/articleshow/90529373.cms

6. Azim Premji University.(2022). Bangalore Covid Impact Survey. Azim Premji University. https://cse.azimpremjiuniversity.edu.in/wp-content/uploads/2022/03/bcis_public-ppt_short.pdf

7. PTI. BBMP allows all business establishments to operate, but with certain conditions. Deccan Herald. 2021.

8. COVID-19 test must for factory workers. (2020, Oct 31). The Hindu. https://www.thehindu.com/news/national/karnataka/COVID-19-test-must-for-factory-workers/article32987064.ece

9. ICMR. Guidance to enhance availability of COVID-19 testing kits and newer innovative testing solutions in India India: Indian Council of Medical Research (ICMR); 2021 [Available from: https://www.icmr.gov.in/pdf/covid/kits/Guidance_COVID_testing_commodities_18062021.pdf.

10. McElfish PA, Purvis R, James LP, Willis DE, Andersen JA. Perceived Barriers to COVID-19 Testing. Int J Environ Res Public Health. 2021 Feb 25;18(5):2278. doi: 10.3390/ijerph18052278. PMID: 33668958; PMCID: PMC7956381.

11. Barnagarwala.T. (2021,Aug 10).In India, over 40% districts still don’t have a single RT-PCR lab for COVID-19 testing. Scroll. in. https://scroll.in/article/1002314/in-india-over-40-districts-still-dont-have-a-single-rt-pcr-lab-for-COVID-19-testing

12. covid 19 information work places-rapid antigen testing.(2022*, 13 Jan).* in safe work australia. retrieved from https://covid19.swa.gov.au/COVID-19-information-workplaces/industry-information/building-and-construction/rapid-antigen

13. Rosella LC, Agrawal A, Gans J, Goldfarb A, Sennik S, Stein J. Large-scale implementation of rapid antigen testing system for COVID-19 in workplaces. Sci Adv. 2022 Feb 25;8(8):eabm3608. doi: 10.1126/sciadv.abm3608. Epub 2022 Feb 25. PMID: 35213224; PMCID: PMC8880770.

14. Goggolidou P, Hodges-Mameletzis I, Purewal S, Karakoula A, Warr T. Self-Testing as an Invaluable Tool in Fighting the COVID-19 Pandemic. J Prim Care Community Health. 2021 Jan-Dec;12:21501327211047782. doi: 10.1177/21501327211047782. PMID: 34583571; PMCID: PMC8485257.

15. Napierala Mavedzenge S, Baggaley R, Corbett EL. A review of self-testing for HIV: research and policy priorities in a new era of HIV prevention. Clin Infect Dis. 2013 Jul;57(1):126–38. doi: 10.1093/cid/cit156. Epub 2013 Mar 13. PMID: 23487385; PMCID: PMC3669524.

16. Hamilton A, Thompson N, Choko AT, Hlongwa M, Jolly P, Korte JE, et al. HIV Self-Testing Uptake and Intervention Strategies Among Men in Sub-Saharan Africa: A Systematic Review. Frontiers in Public Health. 2021;9.

17. World Health Organization. (2019) WHO recommends HIV self-testing – evidence update and considerations for success. World Health Organization.https://www.who.int/publications/i/item/WHO-CDS-HIV-19.36

18. World Health Organization. (2021). Recommendations and guidance on hepatitis C virus self-testing. World Health Organization. https://apps.who.int/iris/handle/10665/342803.

19. World Health Organization. (2022). Use of SARS-CoV-2 antigen-detection rapid diagnostic tests for COVID-19 self-testing. World Health Organization. https://www.who.int/publications/i/item/WHO-2019-nCoV-Ag-RDTs-Self_testing-2022.1

20. Shahmanesh M, Mthiyane TN, Herbsst C, Neuman M, Adeagbo O, Mee P, et al. Effect of peer-distributed HIV self-test kits on demand for biomedical HIV prevention in rural KwaZulu-Natal, South Africa: a three-armed cluster-randomised trial comparing social networks versus direct delivery. BMJ Global Health. 2021;6(Suppl 4):e004574.

21. Okoboi S, Lazarus O, Castelnuovo B, Nanfuka M, Kambugu A, Mujugira A, et al. Peer distribution of HIV self-test kits to men who have sex with men to identify undiagnosed HIV infection in Uganda: A pilot study. PLoS One. 2020;15(1):e0227741.

22. Sibanda, Euphemia, Choko, Augustine Talumba, Watadzaushe, Connie, Mukoka, Madalo, Kumwenda, Moses., et al. (2022). Use of SARS-CoV-2 antigen-detection rapid diagnostic tests for COVID-19 self-testing: interim guidance, 9 March 2022: web annex B: COVID-19 self-testing using antigen rapid diagnostic tests: feasibility evaluation among health-care workers and general population in Malawi and Zimbabwe. World Health Organization. https://apps.who.int/iris/handle/10665/352345. License: CC BY-NC-SA 3.0 IGO

23. WHO. WHO Emergency Use Assessment Coronavirus disease (COVID-19) IVDs. Geneva: World Health Organization (WHO); 2021. Contract No.: EUL-0587-032-00.

24. Reipold, E.I., Farahat, A., Elbeeh, A. et al. Usability and acceptability of self-testing for hepatitis C virus infection among the general population in the Nile Delta region of Egypt. BMC Public Health 21, 1188 (2021).

25. Wanat M, Logan M, Hirst JA, et al Perceptions on undertaking regular asymptomatic self-testing for COVID-19 using lateral flow tests: a qualitative study of university students and staff BMJ Open 2021;11:e053850. doi: 10.1136/bmjopen-2021-053850.

26. Wachinger J, Schirmer M, Täuber N, et al Experiences with opt-in, at-home screening for SARS-CoV-2 at a primary school in Germany: an implementation study BMJ Paediatrics Open 2021;5:e001262. doi: 10.1136/bmjpo-2021-001262.

27. Lamb, Georgia, et al. ‘Real-World Evaluation of COVID-19 Lateral Flow Device (LFD) Mass-Testing in Healthcare Workers at a London Hospital; a Prospective Cohort Analysis’. Journal of Infection, vol. 83, no. 4, Oct. 2021, pp. 452–57. *DOI.org (Crossref)*, 10.1016/j.jinf.2021.07.038.

28. Goggolidou P, Hodges-Mameletzis I, Purewal S, Karakoula A, Warr T. Self-Testing as an Invaluable Tool in Fighting the COVID-19 Pandemic. J Prim Care Community Health. 2021 Jan-Dec;12:21501327211047782. doi: 10.1177/21501327211047782. PMID: 34583571; PMCID: PMC8485257.

29. Nguyen, N., Lane, B., Lee, S., Gorman, S. L., Wu, Y., Li, A., Lu, H., Elhadad, N., Yin, M., & Meyers, K. (2022). A mixed methods study evaluating acceptability of a daily COVID-19 testing regimen with a mobile-app connected, at-home, rapid antigen test: Implications for current and future pandemics. PloS one, 17(8), e0267766. 10.1371/journal.pone.0267766

30. Laura C. Rosella et al., Large-scale implementation of rapid antigen testing system for COVID-19 in workplaces. Sci. Adv.8,eabm3608(2022).DOI:10.1126/sciadv.abm3608 Export citation

31. Abu Hassan, M. R., Addul Rahman, S., Chan, H. K., Zulkifli, N. F., Sem, X., Marbán-Castro, E., Redzuan, S., Abdul Aziz, A., Del Rey, P., & Shilton, S. (2023). IMPLEMENTATION PILOT OF A COVID-19 SELF-TESTING MODEL FOR EMPLOYEES OF MANUFACTURING INDUSTRIES AND THEIR HOUSEHOLD MEMBERS IN KEDAH STATE, MALAYSIA. Poster Presentation at 1st Australasian Conference on Point of Care Testing for Infectious Diseases (POC 2023) taking place at the Sheraton Grand Sydney Hyde Park Tuesday 14 - Wednesday 15 March 2023.

32. M. Bresser, R.M. Erhardt, K. Shanaube, M. Simwinga, P.A. Mahlatsi, J. Belus, A. Schaap, A. Amstutz, T. Gachie, T.R. Glass, B. Kangolo, M.J. ‘Mota, S. Floyd, B. Katende, E. Klinkenberg, H. Ayles, K. Reither, M. Ruperez. Evaluation of COVID-19 Ag-RDTs self-testing in Lesotho and Zambia medRxiv 2022.12.21.22283827; 10.1101/2022.12.21.22283827

33. Stohr, J. J. J. M., Zwart, V. F., Goderski, G., Meijer, A., Nagel-Imming, C. R. S., Kluytmans-van den Bergh, M. F. Q., Pas, S. D., van den Oetelaar, F., Hellwich, M., Gan, K. H., Rietveld, A., Verweij, J. J., Murk, J. L., van den Bijllaardt, W., & Kluytmans, J. A. J. W. (2022). Self-testing for the detection of SARS-CoV-2 infection with rapid antigen tests for people with suspected COVID-19 in the community. Clinical microbiology and infection : the official publication of the European Society of Clinical Microbiology and Infectious Diseases, 28(5), 695–700. 10.1016/j.cmi.2021.07.039

34. Simwinga M, Kumwenda MK, Dacombe RJ, Kayira L, Muzumara A, Johnson CC, Indravudh P, Sibanda EL, Nyirenda L, Hatzold K, Corbett EL, Ayles H, Taegtmeyer M. Ability to understand and correctly follow HIV self-test kit instructions for use: applying the cognitive interview technique in Malawi and Zambia. J Int AIDS Soc. 2019 Mar;22 Suppl 1(Suppl Suppl 1):e25253. doi: 10.1002/jia2.25253. PMID: 30907496; PMCID: PMC6432102

35. Papenburg J, Campbell JR, Caya C, et al. Adequacy of Serial Self-performed SARS-CoV-2 Rapid Antigen Detection Testing for Longitudinal Mass Screening in the Workplace. JAMA Netw Open. 2022;5(5):e2210559. doi:10.1001/jamanetworkopen.2022.10559

36. Peck, R.B., Lim, J.M., van Rooyen, H. et al. What Should the Ideal HIV Self-Test Look Like? A Usability Study of Test Prototypes in Unsupervised HIV Self-Testing in Kenya, Malawi, and South Africa. AIDS Behav 18 (Suppl 4), 422–432 (2014). 10.1007/s10461-014-0818-8

37. Majam M, Mazzola L, Rhagnath N, Lalla-Edward ST, Mahomed R, Venter WDF, Fischer AE. Usability assessment of seven HIV self-test devices conducted with lay-users in Johannesburg, South Africa. PLoS One. 2020 Jan 14;15(1):e0227198. doi: 10.1371/journal.pone.0227198. PMID: 31935228; PMCID: PMC6959591.

38. Muwanguzi, P.A., Bollinger, R.C., Ray, S.C. et al. Drivers and barriers to workplace-based HIV self-testing among high-risk men in Uganda: a qualitative study. BMC Public Health 21, 1002 (2021). 10.1186/s12889-021-11041-y

39. Peck, R.B., Lim, J.M., van Rooyen, H. et al. What Should the Ideal HIV Self-Test Look Like? A Usability Study of Test Prototypes in Unsupervised HIV Self-Testing in Kenya, Malawi, and South Africa. AIDS Behav 18 (Suppl 4), 422–432 (2014). 10.1007/s10461-014-0818-8

40. Atchison C, Pristerà P, Cooper E, Papageorgiou V, Redd R, Piggin M, Flower B, Fontana G, Satkunarajah S, Ashrafian H, Lawrence-Jones A, Naar L, Chigwende J, Gibbard S, Riley S, Darzi A, Elliott P, Ashby D, Barclay W, Cooke GS, Ward H. Usability and Acceptability of Home-based Self-testing for Severe Acute Respiratory Syndrome Coronavirus 2 (SARS-CoV-2) Antibodies for Population Surveillance. Clin Infect Dis. 2021 May 4;72(9):e384–e393. doi: 10.1093/cid/ciaa1178. PMID: 32785665; PMCID: PMC7454392.

41. Majam M, Mazzola L, Rhagnath N, Lalla-Edward ST, Mahomed R, Venter WDF, et al. (2020) Usability assessment of seven HIV self-test devices conducted with lay-users in Johannesburg, South Africa. PLoS ONE 15(1): e0227198. 10.1371/journal.pone.0227198

42. Pai N, Esmail A, Saha Chaudhuri P, et al. Impact of a personalised, digital, HIV self-testing app-based program on linkages and new infections in the township populations of South Africa. BMJ Global Health 2021;6:e006032.

43. McGuire, M., de Waal, A., Karellis, A., Janssen, R., Engel, N., Sampath, R., Carmona, S., Zwerling, A. A., Suarez, M. F., & Pai, N. P. (2021). HIV self-testing with digital supports as the new paradigm: A systematic review of Global Evidence (2010–2021). EClinicalMedicine, 39, 101059. 10.1016/j.eclinm.2021.101059

44. Rao, A., Patil, S., Kulkarni, P.P. et al. Acceptability of HIV oral self-test among truck drivers and youths: a qualitative investigation from Pune, Maharashtra. BMC Public Health 21, 1931 (2021). 10.1186/s12889-021-11963-7

45. WHO guideline on self-care interventions for health and well-being, 2022 revision. (2022, June 27). WHO Guideline on Self-care Interventions for Health and Well-being, 2022 Revision. https://www.who.int/publications/i/item/9789240052192

46. COST-EFFECTIVENESS OF WORKPLACE COVID-19 SELF-TESTING: A MATHEMATICAL MODELING STUDY - CROI Conference. (2023, March 4). CROI Conference -.https://www.croiconference.org/abstract/cost-effectiveness-of-workplace-COVID-19-self-testing-a-mathematical-modeling-study/

